# Integrative genetic and genomic networks identify microRNA associated with COPD and ILD

**DOI:** 10.1101/2022.08.07.22278496

**Authors:** Ana B. Pavel, Carly Garrison, Lingqi Luo, Gang Liu, Daniel Taub, Ji Xiao, Brenda Juan-Guardela, John Tedrow, Yuriy O. Alekseyev, Ivana V. Yang, Mark W. Geraci, Frank Sciurba, David A. Schwartz, Naftali Kaminski, Jennifer Beane, Avrum Spira, Marc E. Lenburg, Joshua D. Campbell

## Abstract

Chronic obstructive pulmonary disease (COPD) and interstitial lung disease (ILD) are clinically and molecularly heterogeneous diseases. We utilized clustering and integrative network analyses to elucidate roles for microRNAs (miRNAs) and miRNA isoforms (isomiRs) in COPD and ILD pathogenesis. Short RNA sequencing was performed on 351 lung tissue samples of COPD (n=145), ILD (n=144) and controls (n=64). Five distinct subclusters of samples were identified including 1 COPD-predominant cluster and 2 ILD-predominant clusters which associated with different clinical measurements of disease severity. Utilizing 262 samples with gene expression and SNP microarrays, we built disease-specific genetic and expression networks to predict key miRNA regulators of gene expression. Members of miR-449/34 family, known to promote airway differentiation by repressing the Notch pathway, were among the top connected miRNAs in both COPD and ILD networks. Genes associated with miR-449/34 members in the disease networks were enriched among genes that increase in expression with airway differentiation at an air-liquid interface. A highly expressed isomiR containing a novel seed sequence was identified at the miR-34c-5p locus. 47% of the anticorrelated predicted targets for this isomiR were distinct from the canonical seed sequence for miR-34c-5p. Overexpression of the canonical miR-34c-5p and the miR-34c-5p isomiR with an alternative seed sequence down-regulated NOTCH1 and NOTCH4. However, only overexpression of the isomiR down-regulated genes involved in Ras signaling such as CRKL and GRB2. Overall, these findings elucidate molecular heterogeneity inherent across COPD and ILD patients and further suggest roles for miR-34c in regulating disease-associated gene-expression.

## INTRODUCTION

Complex chronic lung diseases arise from heterogeneous molecular processes and are influenced by multiple factors including exposure to toxins and genetic susceptibility. Chronic obstructive pulmonary disease (COPD) is a progressive lung disease and the fourth leading cause of death worldwide,^1^ with an incidence of 2.8 cases per 1,000 persons per year^2^. Patients with COPD suffer from breathing difficulty, wheezing, excess mucus production, and chronic cough. Although biological processes, such as chronic inflammation, apoptosis, and oxidative stress, have been implicated in COPD pathogenesis, knowledge of the key molecular drivers of this disease remains limited^3^. Interstitial lung disease (ILD) is a collection of chronic lung diseases characterized by fibrosis or inflammation of the alveolar tissue in the lung parenchyma^4^. One of the most common subtypes of ILD, idiopathic pulmonary fibrosis (IPF), has an incidence of 6.8-8.8 per 100,000 persons per year^5^ with a median survival from diagnosis of 3–5 years^6–8^.

MicroRNAs (miRNAs) are short RNA transcripts about 20-23 nucleotides long that can modulate expression levels or translation rates of specific mRNA targets via sequence-specific binding to their 3’ UTR^9^. MicroRNAs are involved in a wide variety of developmental processes and aberrant activity of miRNAs can also contribute to disease pathogenesis^10^. Previous studies have performed transcriptomic profiling of affected lung tissue to understand the molecular processes associated with complex lung diseases such as COPD and ILD^11–13^. Additional studies sought to identify microRNA (miRNA) expression profiles associated with the presence of disease to gain insights into the regulation of aberrant gene expression^14–16^. Despite the information gained from these initial studies, larger sample sets are needed to identify novel molecular subtypes of disease and more data modalities need to be measured on each sample to perform integrative network analyses.

Network approaches that integrate multiple data types have been used extensively to study complex diseases^17^. Integrative genetic and genomic network approaches have been used to identify molecular drivers of late-onset Alzheimer’s disease and breast cancer risk^18,19^. Integrative analysis of DNA methylation and gene expression has identified key regulators in the setting of COPD^20^. Several computational approaches have been applied to infer causality from biological data, including Bayesian networks,^21–23^ factor graphs^18,19^ and ridge and least absolute shrinkage and selection operator^24^. Statistical framework such as the Causlity Inference Test (CIT), can be used to infer mediators of genetic or epigenetic factors associated with quantitative traits^25^. The CIT has also been used to characterize the role of microRNAs (miRNAs) in gene regulatory networks^26^ and can be applied in settings where profiling of miRNA expression, mRNA expression, and genetic or epigenetic variation have been captured on the same samples.

Previous miRNA studies in COPD and ILD have relied primarily on microarray technology for quantifying expression. Microarrays only allow for the profiling of a canonical miRNA sequences. With the advent of small RNA sequencing, additional variation in miRNA sequences have been observed including variation on the 5’ end of mature miRNAs^27^. Variation on the 5’ end of a miRNA produces a different seed sequence. The seed sequence is the primary feature for determining the binding specificity of the miRNA to the 3’ UTR of mRNA transcripts. These noncanonical miRNAs, often called isomiRs, can have alternative functional roles compared to the canonical miRNA sequence at that locus due to the targeting of distinct mRNAs^28^. Despite the potentially important role of isomiRs in regulating gene expression, the expression patterns of isomiRs have not been well-characterized in tissues from subjects with chronic lung disease.

In order to characterize molecular heterogeneity in chronic lung disease and predict key regulators of gene expression, we profiled miRNA expression via small-RNA sequencing from a large number of samples from the Lung Genome Research Consortium (LGRC). Unsupervised clustering revealed subgroups of subjects with distinct clinical and molecular characteristics. Using the CIT, we developed integrative networks and found increased connectivity in the disease cohorts for miRNAs involved in airway differentiation and ciliogenesis. Finally, we identified and characterized a 5’ isomiR of miR-34c-5p with putative roles in the regulation of Ras pathway members. Overall, these analyses provide a comprehensive view of miRNA expression patterns COPD and ILD and elucidate the potential roles of these miRNA in regulating biological pathways within the lung.

## METHODS

### High-throughput sequencing of small RNA

RNA was obtained from the National Heart, Lung, and Blood Institute–sponsored Lung Tissue Research Consortium (LTRC)^11,13^. Tissue samples from the LTRC are labeled with IDs that cannot be used to identify to the study subjects. 45 samples were prepared with Small RNA Sample Prep Kit v1.5 (Illumina) and sequenced on the Genome Analyzer IIx (Illumina) according to the manufacturer’s protocol. Multiplexed small RNA sequencing was conducted on the Illumina HiSeq 2000 for 320 lung tissue samples. Briefly, one microgram of total RNA from each sample was used for library preparation with a TruSeq Small RNA Sample Prep Kit (Illumina). RNA adapters were ligated to 3’ and 5’ end of the RNA molecule and the adapter-ligated RNA was reverse transcribed into single-stranded cDNA. The RNA 3’ adapter was specifically designed to target miRNAs and other small RNAs that have a 3’ hydroxyl group resulting from enzymatic cleavage by Dicer or other RNA processing enzymes. The cDNA was then PCR amplified using a common primer and a primer containing one of 10 index sequences. The introduction of the six-base index tag at the PCR step allowed multiplexed sequencing of different samples in a single lane of a flowcell. Ten individual PCR-enriched cDNA libraries with unique indices in equal amount were pooled and gel purified together. A 0.5% PhiX spike-in was also added in all lanes for quality control. Each library was hybridized to one lane of the 8-lane single-read flowcell on a cBot Cluster Generation System (Illumina) using TruSeq Single-Read Cluster Kit (Illumina). The clustered flowcell was loaded onto HiSeq 2000 sequencer for a multiplexed sequencing run that consists of a standard 36-cycle sequencing read with the addition of a 7-cycle index read.

### miRNA alignment and quality control

To estimate miRNA expression we used a small RNA sequencing pipeline previously described^29^. Briefly, the 3’ adapter sequence was trimmed using the FASTX toolkit. Reads longer than 15 nt were aligned to hg19 using Bowtie v0.12.7^30^ allowing up to one mismatch and up to 10 genomic locations. miRNA expression was quantified by counting the number of reads aligning to mature miRNA loci (miRBase v20) using Bedtools v2.9.0.^31,32^ For quality control, we examined the distribution of read lengths for each sample after trimming to ensure that the sequences we observed were of the proper length for miRNA. 13 of 365 samples clustered differently than the rest of the samples based on the read length distribution and were excluded from subsequent analyses (**Supplementary Figure 1**). One additional sample was excluded as a duplicate leaving 351 samples for expression analysis (**Table 1**).

**Table 1.**
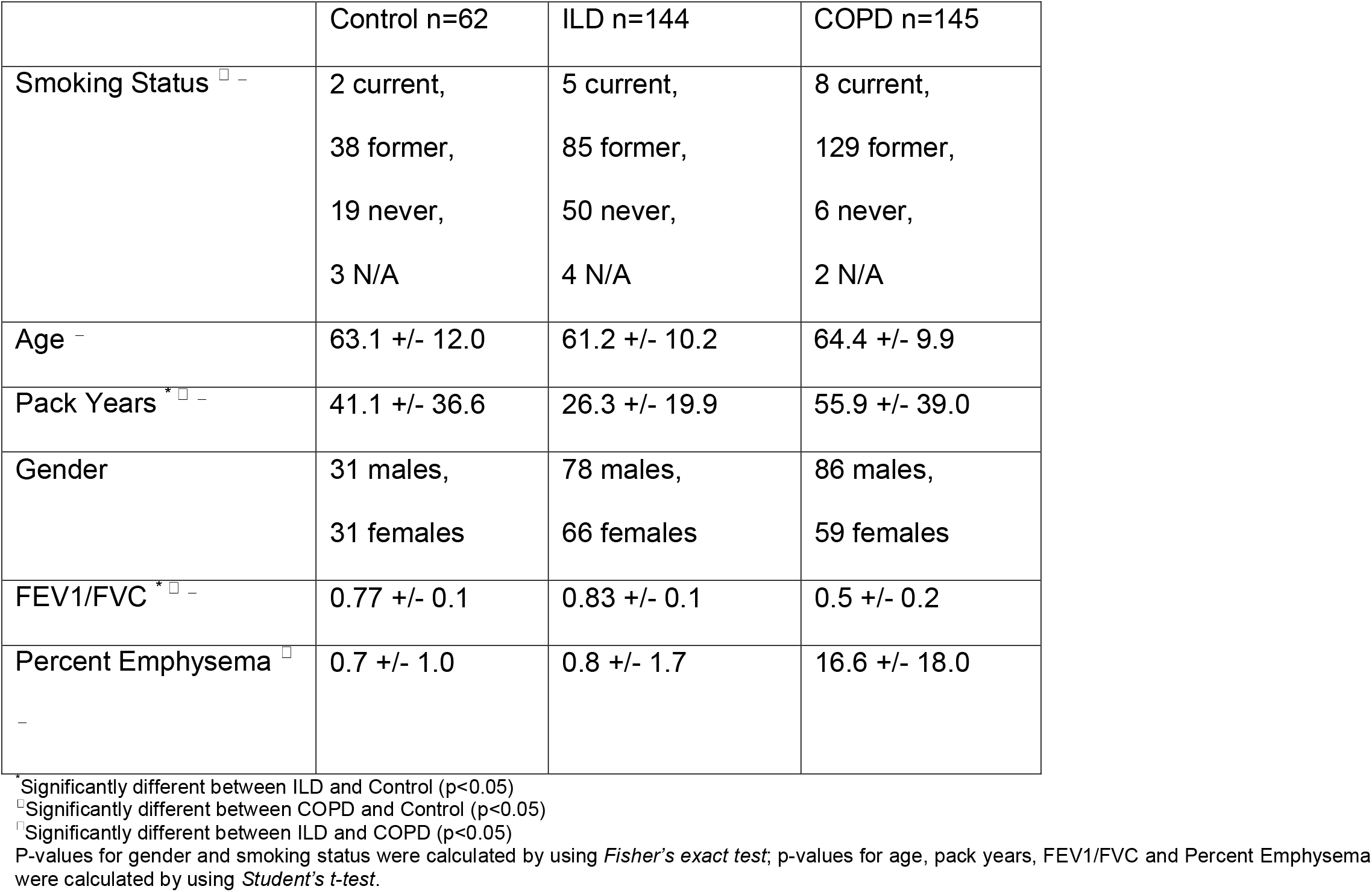
Sample demographics.

### Differential expression

To identify miRNAs associated with disease a generalized negative binomial model (*glm*.*nb*, MASS R package) was applied to each miRNA. The count of each miRNA was used as the response variable and sequencing read depth, sequencing protocol, smoking status, age, gender, as well as COPD status and ILD status were used as predictor variables. The significance of the associations were assessed by performing an ANOVA^33^ between a model with both the COPD and ILD terms and a second model without the either disease term. P-values were adjusted with the Benjamini-Hochberg false discovery rate^34^ (FDR). MiRNAs were considered differentially expressed if they had an FDR q-value < 0.1 and the absolute value of the coefficient for either the COPD or ILD term was greater than 0.22, corresponding to a Fold Change (FC) of at least 1.25.

### Consensus Clustering

For clustering and display in heatmaps, miRNA counts within each sample were normalized to RPM values by adding a pseudocount of one to each miRNA, dividing by the total number of reads that aligned to all miRNA loci within that sample, multiplying by 1 × 10^6^, and then applying a log_2_ transformation.^29^ The batch effects of the two sequencing protocols were removed by Combat.^35^ Groups of miRNAs or samples were identified using consensus clustering^36^ (*ConsensusClusterPlus* R package) on the normalized and batch-corrected miRNA expression data. Sample clusters were assessed for enrichment of disease samples by using two logistic regression models where disease status (either COPD or ILD) was the response and sequencing read depth, sequencing protocol, smoking status, age, gender, and cluster status were dependent variables. Sample clusters were also associated with clinical variables of disease severity including DLCO (diffusing capacity of the lungs for carbon monoxide), FEV1/FVC ratio (forced expiratory volume 1 / forced vital capacity), FEV1 percent predicted, percent emphysema, and BODE score (i.e., a measure of the degree of obstruction, dyspnea, and exercise capacity). Linear models were fit where each clinical phenotype was the response variable and sequencing read depth, sequencing protocol, smoking status, age, gender, and cluster status were dependent variables. Two separate models were fit for ILD and COPD patients.

### Building disease specific networks

We utilized a subset of 262 lung tissue samples with miRNA expression profiled by sequencing, as well as an Agilent gene expression microarray and Affymetrix SNP chip. We first identified all genes and miRNAs associated with a SNP (i.e. eQTL) by ANOVA while correcting for age, gender, smoking status, and population structure (p<0.0005) using the *MatrixEQTL* package.^37^ Next, we built integrative networks within the COPD, ILD, and control patients using the causality inference test (CIT).^25^ This test is a previously established method for predicting SNP-miRNA-mRNA triplets where the SNP is modulating the expression of the miRNA and the miRNA is modulating the expression of the gene.^25^ CIT assesses the hypothesis that a potential mediator between a genetic variable and an outcome variable is potentially causal for that outcome. Causal and reactive models are defined as series of conditions of associations between the three variables, corresponding to SNP, microRNA and mRNA nodes. The significance of the test is computed for both the causal and reactive models. If the causal p-value is lower than 0.05 and the reactive higher than 0.05 then the causal relationship is indicated. If both p-values are greater than 0.05 then the call is independent, and if both p-values are lower than 0.05, then causality cannot be inferred. We select those SNP-miRNA-mRNA triplets where the SNP-mRNA relationship is defined by a miRNA mediator and did not examine triplets where the SNP is not associated with the miRNA. The number of mRNA predicted to be regulated by each miRNA was compared between control and disease networks. The genes found to be regulated by the top differentially connected miRNAs were examined by GSVA^38^ and GSEA^39^ in an independent dataset examining gene expression patterns associated with differentiation of airway epithelium at an air-liquid interface (ALI)^40^.

### Validation by qRT-PCR

To measure the expression of miR-34a-5p, miR-34b-5p and miR-34c-5p, 10 ng of total RNA was used in a Taqman MiRNA Assay (Life Technologies, Catalog #4427975, ID #000426, 000427, 000428, Carlsbad, CA) as per manufacturer’s protocol and the results were normalized to RNU44 expression (Life Technologies, Catalog #4427975, ID #001094, Carlsbad, CA). To measure the expression of *RALA, GRB2, CRK, CRKL, GRAP, RHOA, RHOC, EGF, ARAP2, NOTCH4* and *NOTCH1*, 500 ng of total RNA was reverse transcribed using RT2 First Strand Kits (Qiagen, Catalog #330401, Valencia, CA) according to the manufacturer’s protocol. cDNA product was added to SYBR Green qPCR Mastermix (Qiagen, Catalog #330523, Valencia, CA) and the appropriate primer (Qiagen, Catalog #PPH07458A, PPH00714C, PPH00731A, PPH01982A, PPH13173A, PPH00305G, PPH01089E, PPH00137B, PPH20012A, PPH06021F, PPH00526C, Valencia, CA). Data was normalized to the expression of UBC (Qiagen, Catalog #PPH00223F, Valencia, CA) and analyzed using the comparative CT method.

### Quantification and transfection of isomiRs

IsomiRs were identified within each canonical miRNA locus by grouping reads with the same 5’ start position. Targetscan v6.0^41^ was used to predict mRNA targets for each canonical and isomiR seed. IMR90 cells and HBEpCs were transiently transfected with hsa-miR-34c-5p miRIDIAN miRNA mimic (Dharmacon, Catalog #C-300655-03-0020, Lafayette, CO), a custom miR-34c 5’ isomiR miRIDIAN miRNA mimic (Dharmacon, Lafayette, CO) or miRIDIAN miRNA mimic Negative Control #1 (Dharmacon, Catalog #C-001000-01, Lafayette, CO). IMR90 cell transfection was completed using Lipofectamine RNAiMAX transfection reagent (Life Technologies, Catalog #13778150, Carlsbad, CA) according to the manufactures protocol. Transfection of HBEpCs was done using Cytofect Epithelial Cell Transfection Kit (Cell Applications, Catalog #TF102K, San Diego, CA).

### Data availability

SNP data was provided by the Lung Genomics Research Consortium (LGRC; http://lung-gemomics.org; 1RC2HL101715) using tissue samples and clinical data collected through the Lung Tissue Research Consortium (LTRC; http://www.ltrcpublic.com/). This data is available from dbGaP under the accession phs000624.v1.p1. The microRNA expression datasets generated and analyzed during the current study are available in the Raw and normalized data is available at the Gene Expression Omnibus (GEO) under the accession number GSE201121.

## RESULTS

### Subject cohort

MicroRNA expression was profiled with small-RNA sequencing for 364 lung tissue samples collected by the Lung Tissue Research Consortium. Thirteen samples with low quality were removed (**Methods; Supplementary Table 1**) resulting in 351 samples for downstream analyses including 145 subjects with COPD, 144 subjects with ILD, 62 Controls (**Table 1, Supplementary Table 2**). Controls were mostly derived from tissue adjacent normal to cancer as previously described^11^. Subjects with COPD had a significantly higher proportion of former smokers, higher Pack Years, lower FEV1/FVC ratios, and higher Percent Emphysema compared to Control subjects. Subjects with ILD had significantly lower Pack Years and higher FEV1/FVC ratios compared to Control subjects. Compared to individuals with ILD, COPD subjects had a significantly higher proportion of former smokers, higher Age, higher FEV1/FVC ratios, and higher Percent Emphysema.

### Unsupervised clustering identifies novel subgroups associated with clinical phenotypes

693 of 2104 mature miRNAs were detected with 2 counts in at least 50% of samples. The expression profiles of 255 miRNAs were significantly associated with the presence of disease (ANOVA FDR q-value < 0.10 and fold change > 1.25 in either disease group compared to controls; **Figure 1A; Supplementary Table 2**). Five clusters of samples (S1-5) and 4 clusters of miRNA (M1-4) were determined by Consensus Clustering (**Methods; Supplementary Figure 2**). The majority of control samples (52%) were found in S1 (**Figure 1B**). Clusters S2, S4, and S5 had significantly more ILD samples compared to S1 (p<0.05). While the fractions of COPD samples in clusters S2-S5 were not significantly higher compared to cluster S1 (p>0.05), sample cluster S3 contained the highest proportion of COPD cases (53%).

**Figure 1.**
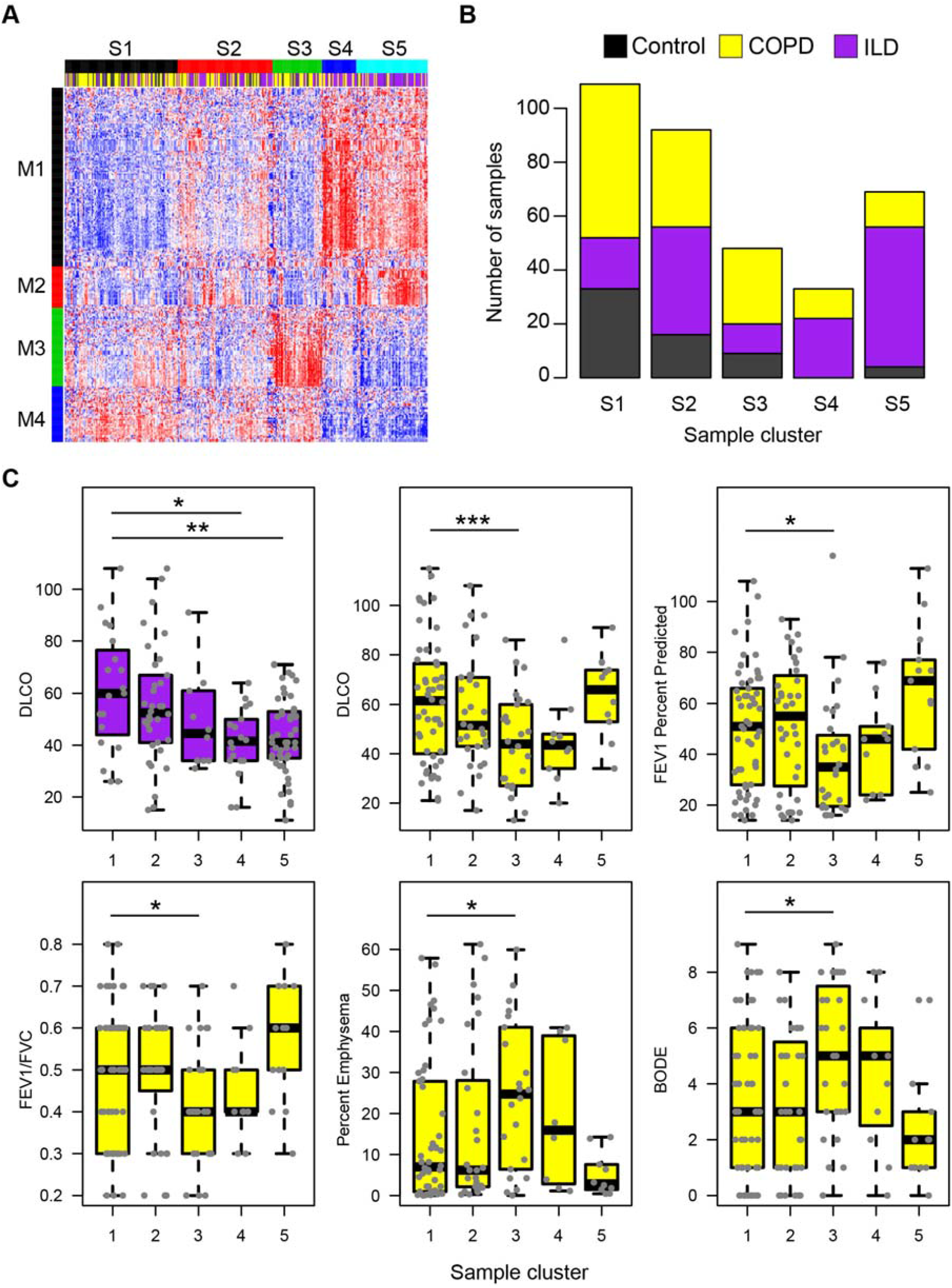
Heterogeneity of miRNA expression profiles associated with COPD or ILD. **A**. The expression profiles of 255 miRNAs were significantly associated with the presence of disease (ANOVA FDR q-value < 0.10 and fold change > 1.25 in either disease group compared to controls). Consensus clustering was used to identify 5 distinct samples clusters and 4 distinct miRNA clusters. **B**. Stacked barplots display the proportion of disease and control samples within each sample cluster. The majority of control samples (53%) fell into cluster S1. Clusters S2, S4, and S5 were enriched with ILD patients compared to cluster S1. **C**. ILD samples in clusters S4 and S5 has significantly lower DLCO compared to ILD samples in cluster S1 (p < 0.05). COPD samples in cluster S3 had significantly lower DLCO, FEV1 percent predicted, FEV1/FVC ratios and significantly higher percent emphysema and BODE scores compared to COPD samples in cluster S1. Asterisks indicate significance: * p < 0.05; ** p < 0.01; *** p < 0.001.

The ILD patients in clusters S4 and S5 had significantly lower DLCO (diffusing capacity of the lung for carbon monoxide) compared to ILD patients in cluster 1 (p<0.05; **Figure 1C**). Clusters S4 and S5 had an up-regulation in the expression of miRNAs from M1 and a down-regulation of miRNAs in M4. Sixty of the 125 miRNAs in M1 were regionally co-located on chromosome 14q32 and were previously reported to be up-regulated in IPF^16^. Additional miRNAs in M1, including miR-21-5p/3p, miR-199a-3p, and miR-155-5p, have also been implicated in the ILD subtype IPF^14,42,43^. MiR-199a-5p was also associated with ILD status (p = 0.002) but did not pass our fold change cutoff^42^. The S4 and S5 clusters showed down-regulation in the expression of miRNAs from cluster M4. Several of the miRNAs in the M4 cluster were a part of in the miR-30 family, including miR-30a-5p/3p, miR-30d-5p/3p, miR-30b-5p, and miR-30c-2-3p. The two ILD-associated sample clusters S4 and S5 with more severe disease could be distinguished by higher levels of miRNA cluster M2. M2 contained many miRNAs that are major regulators of airway differentiation and ciliogenesis including miR-34b-5p/3p, miR-34c-5p/3p miR-449a, miR-449b-5p, miR-449c-5p, and miR-4423-5p^44,45^. Other studies have identified two subclasses of IPF that are characterized by differences in expression of ciliary genes^46^. The different patterns of expression of ciliary-related miRNA between clusters S4 and S5 may also be indicative of this subtype. Cluster S2, which was also enriched for ILD samples, had intermediate levels of cluster M1/M2 up-regulation and M4 down-regulation compared to sample clusters S4 and S5, potentially indicating intermediate levels of disease severity.

COPD samples in cluster S3 had significantly lower FEV1 percent predicted (p=0.01), DLCO (p<0.001), and FEV1/FVC (p=0.03), as well as significantly higher BODE score (body-mass index, airflow obstruction, dyspnea, and exercise, p=0.01) and percent emphysema (p=0.01) compared to the COPD samples in cluster S1. Cluster S3 was largely defined by the up-regulation of miRNAs in M3. M3 miRNA included proximal miR-144 and miR-451a cluster on chromosome 17, miR-222-5p and miR-223-5p/-3p on chromosome X, as well as miR-18a and miR-92a-3p from the miR-17-92 polycistronic cluster on chromosome 13. Although not included in our clustering analysis due to the fold change cutoff, other miRNAs in the miR-17-92 polycistronic cluster were also associated with disease status, including miR-17-5p/3p, miR-19b-3p, and miR-20a-5p/3p (FDR q-value < 0.05). Overall, we identified expression patterns of miRNAs that can distinguish unique subsets of patients with COPD and ILD, including patients with more severe clinical phenotypes.

### Comparison of the number of gene and miRNA eQTLs in COPD and ILD

eQTL analysis can reveal insights into specific biological effects that genetic variants have across different tissues or to disease phenotypes.^47^ To compare the numbers of eQTLs between ILD and COPD and the number of eQTL effecting mRNA or miRNA expression in the setting of ILD and COPD, we utilized a subset of 262 lung tissue samples that had data from miRNA sequencing, SNP chips, and mRNA microarrays including 111 COPD, 113 ILD, and 38 Controls (**Supplementary Table 4**). Protein-coding genes and miRNAs associated with a SNP were identified by ANOVA while correcting for age, gender, smoking status, and population structure within ILD, COPD and Control groups (FDR < 0.05; **Table 2**; **Supplementary Figure 3**). The COPD cohort had larger numbers of *trans* eQTLs for both genes and miRNAs compared to the ILD cohort (**Gene:** 110,424 in COPD, 68,050 in ILD; **miRNA:** 557 in COPD, 362 in ILD). In contrast, the ILD cohort had larger numbers of *cis* gene eQTLs and nearly the same number of miRNA eQTLs as the COPD cohort (**Gene:** 8195 in COPD, 10,941 in ILD; **miRNA:** 53 in COPD, 52 in ILD). The proportion of unique genes and miRNAs with at least one *cis* eQTL was similar between COPD and ILD cohorts (***cis* gene:** 6% in COPD, 7% in ILD; ***cis* miRNA**: 2% in COPD, 2% in ILD). However, there was 1.56-fold more *trans* gene eQTLs and 1.53-fold more *trans* miRNA eQTLS in the COPD cohort compared to the ILD cohort. Lastly, the proportion of miRNAs with any eQTL was significantly lower than the proportion of protein-coding genes with any eQTL in both the COPD and ILD cohorts (p < 0.001; Fisher’s exact test; **Table 2**). Overall, these results suggest that there are more *trans* associations contributing to variability in expression in COPD compared to ILD and that miRNAs have fewer proportions of *cis* and *trans* eQTLs than protein-coding genes in both diseases.

**Table 2.** Number of gene and miRNA eQTLs in different diseases.

|  | Interaction Type | COPD (n=111) | ILD (n=113) |
| --- | --- | --- | --- |
| **No. of Gene-SNP eQTLs** | *cis* | 8195 | 10941 |
|  | *trans* | 110424 | 68050 |
| **No. of miRNA-SNP eQTLs** | *cis* | 53 | 52 |
|  | *trans* | 557 | 362 |
| **No. of unique genes with eQTL** | *cis* | 938 (6%) | 1142 (7%) |
|  | *trans* | 6629 (43%) | 4249 (28%) |
| **No. of unique miRNAs with eQTL** | *cis* | 17 (2%) | 14 (2%) |
|  | *trans* | 221 (32%) | 144 (21%) |

### Integrated network analysis implicates the miR-34/449 family in COPD and ILD

Key miRNA regulators of gene expression can more readily be identified in disease states can be more readily identified by “anchoring” expression data with genetic information and building integrative networks.^26^ Genes and miRNAs associated with at least one SNP were included in the network analysis (p < 0.0005). To build integrative networks within each cohort, we leveraged the causality inference test (CIT).^25^ CIT assesses the hypothesis that a potential mediator between an initial random variable and an outcome variable is causal for that outcome. Causal and independent relationships are defined as series of conditions of associations between the three variables, corresponding to SNP, microRNA and mRNA nodes (**Figure 2A**). The number of significant associations obtained at each step of the network construction are presented in **Supplementary Figure 4**. For our study, we focused on relationships where the miRNA is predicted to be the modulator of mRNA expression. A common property of biological networks is that they often display a *scale-free* topology where a few nodes contain the majority of interactions in the network (i.e. the power law).^48,49^ We confirmed that our three networks followed a scale-free topology by observing a strong negative linear relationship between the number of predicted interactions for each microRNA and the frequency of microRNA with a certain number of interactions in log scale (**Figure 2B**). We further examined the miRNAs predicted to interact with the most genes in each network (**Figure 2C, Supplementary Table 5**).

**Figure 2.**
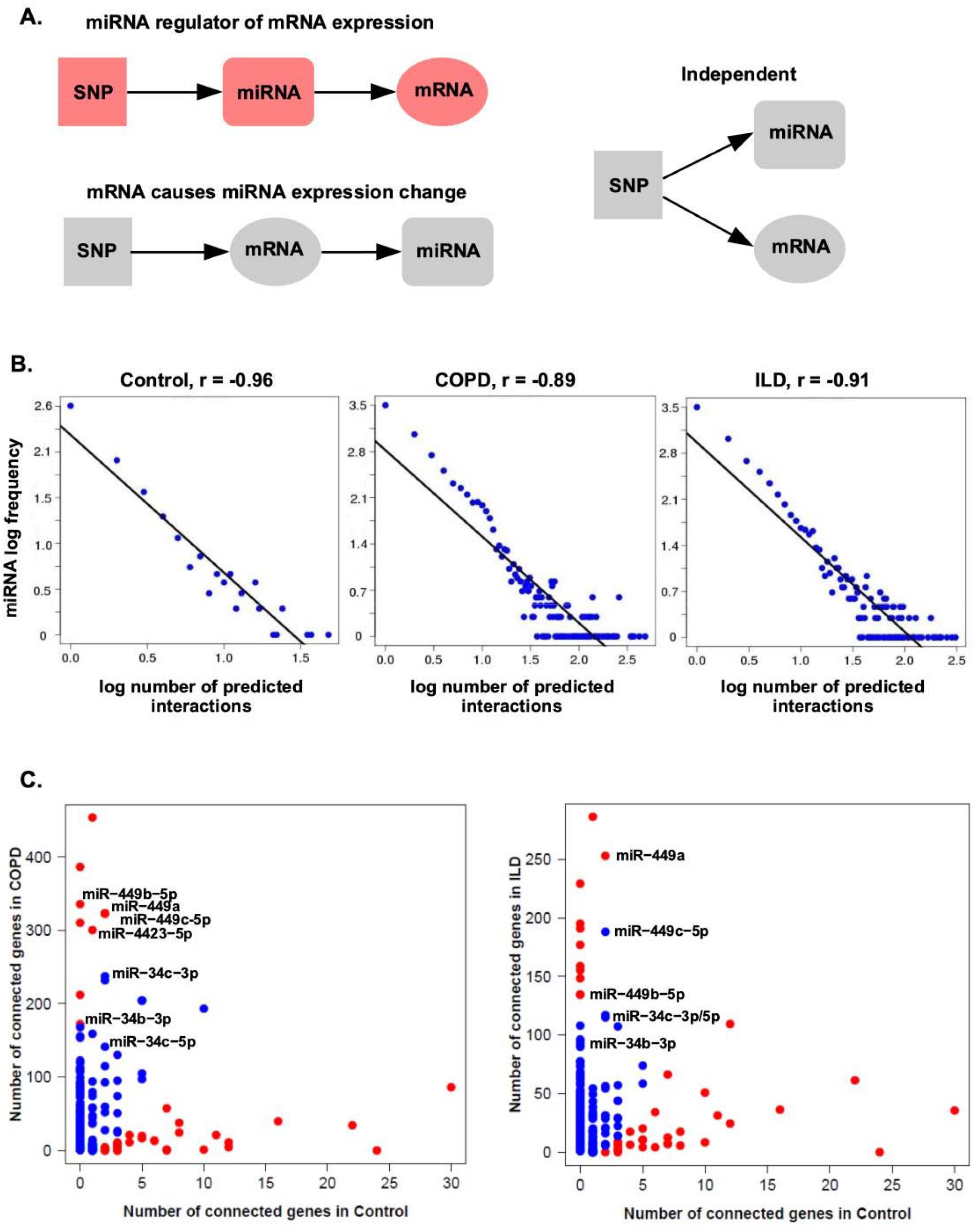
Examining top miRNA in disease-specific integrative networks. **A**. We select those SNP-miRNA-mRNA triplets where the SNP-mRNA relationship is defined by a miRNA mediator; we filter out independent relationships and those triplets where the SNP is not associated with the miRNA. **B**. The CIT networks follow a power law. The negative correlation between the frequency of node degree and the node degree indicates that the networks are *scale-free*. **C**. Number of genes regulated by each miRNA. miR-449/34 family members were found to be among the top 20 differentially connected in COPD and ILD compared to control group. The red dots indicate the significantly differentially connected miRNAs by a Fisher’s exact test (FDR<0.2).

Members of the miR-449 and miR-34 families were found to be among the most connected to genes in COPD and ILD networks (**Figure 2C**), indicating that the miR-449/34 family has a greater impact on gene expression regulation in the disease groups compared to the control group. The miRNAs in the miR449/34 family had larger numbers of associated genes compared to the network in control samples (**Figure 3A**). Members of miR-449/34 family can promote airway differentiation by repressing the Notch pathway^50^. We observed that the union set of genes (n=406) that positively correlated with any of the miRNAs in this family in COPD or ILD was enriched among genes that increase in expression over time when airway basal cells are differentiated at an air-liquid interface (ALI)^40^. Gene enrichment results were significant by both GSVA (p < 0.05; **Figure 3B**) and GSEA (q < 0.001; **Figure 3C**). 75 SNPs in COPD (**Supplementary Table 6**) and 60 SNPs in ILD (**Supplementary Table 6**) were associated with members of the miR-449/34 family using the CIT. Some of these SNPs have been previously found to be associated with asthma, inflammation, cancer and other degenerative diseases in the Genome-Wide Repository of Associations Between SNPs and Phenotypes (GRASP)^51^. Allele frequencies for 10 of these SNPs were also significantly associated with COPD and 4 of them with ILD by a Fisher’s exact test (q<0.25; **Supplementary Figure 5)**.

**Figure 3.**
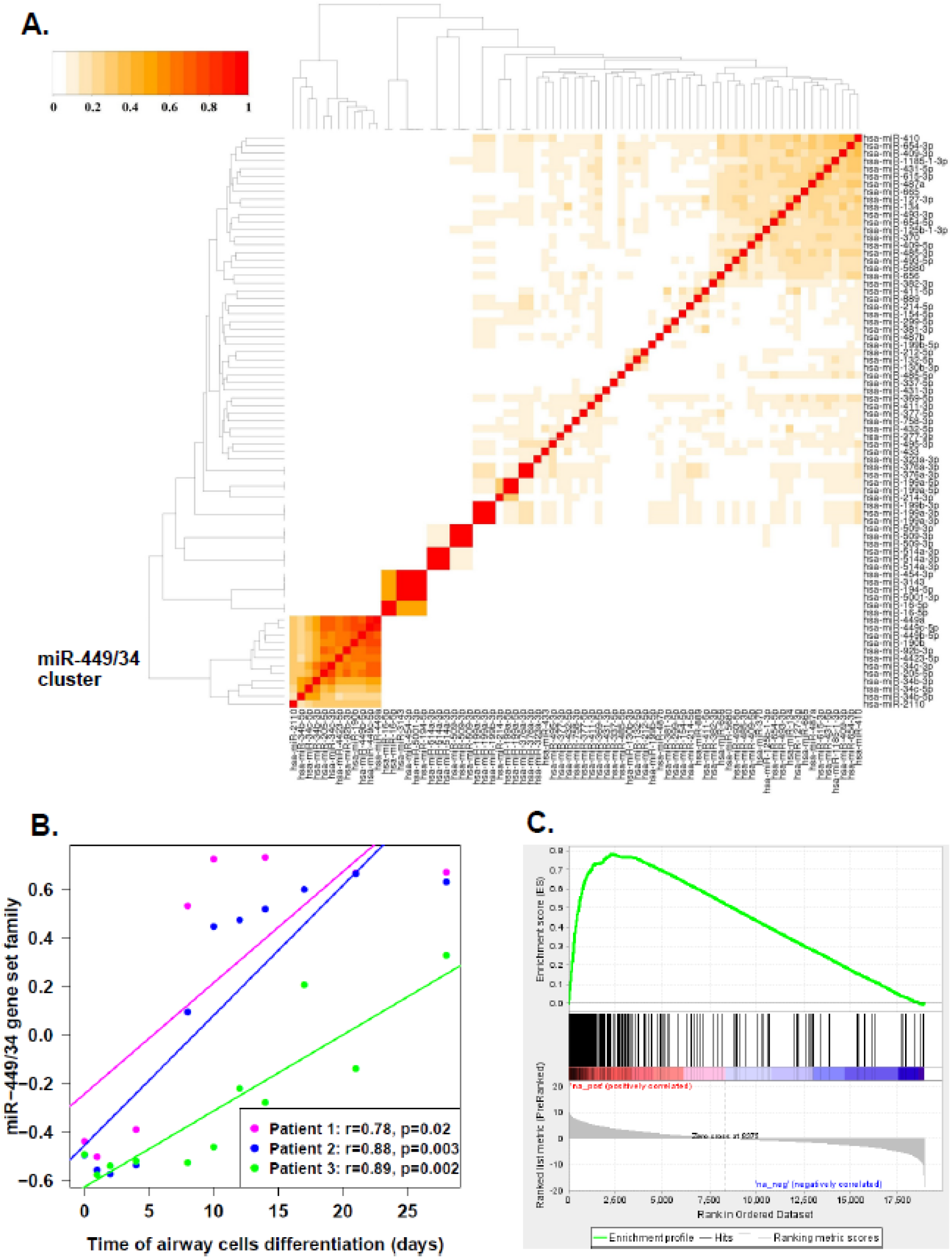
Enrichment of miR-449/34 modules. **A**. Clustering of miRNA modules based on the Jaccard index revealed a group of strongly overlapping miRNA in the miR-449/34 family. **B**. GSVA was used to predict the activity of the miR-449/34 family in a gene-expression dataset of airway epithelial differentiation. The set of genes that positively correlated with miR-449/34 family (406 genes) were enriched among genes that increase in expression with the airway epithelial cells differentiation (p < 0.05; Linear mixed-effects model). **C**. Similarly, enrichment of miR-449/34 gene set family with the airway cells differentiation is shown by GSEA (FDR q-value <0.001).

### The canonical and 5’ isomiR seeds for miR-34c-5p regulate members of distinct signaling pathways

As noted previously, expression for members of the miR-449/34 family were a part of a miRNA module (M2) that could distinguish two distinct groups of ILD patients and were found among the most connected microRNAs in COPD and ILD specific networks. The miR-449/34 family has been shown to promote ciliogenesis by down-regulating anti-differentiation genes such as NOTCH1^52^. The majority of miRNAs in this family share the same seed sequence, GGCAGTG, which is a conserved heptametrical sequence on the 5’ end. Previous studies have revealed that variation in the 5’ end of miRNAs can create a novel seed sequence, which allows the 5’ isomiR to target a distinct set of genes from the canonical seed^27^. Within the miR-34/449 family, we found relatively high expression of isomiRs within the miR-34c-5p locus that contained an alternative seed sequence representing a 1-base shift to the left from the canonical seed, AGGCAGT (**Figure 4A**). 24 of the top 25 sequences from the miR-34c-5p locus were up-regulated in ILD sample compared to the Control samples (**Supplementary Table 8**), demonstrating that the majority of sequences follow the same expression pattern across samples with respect to disease status.

**Figure 4.**
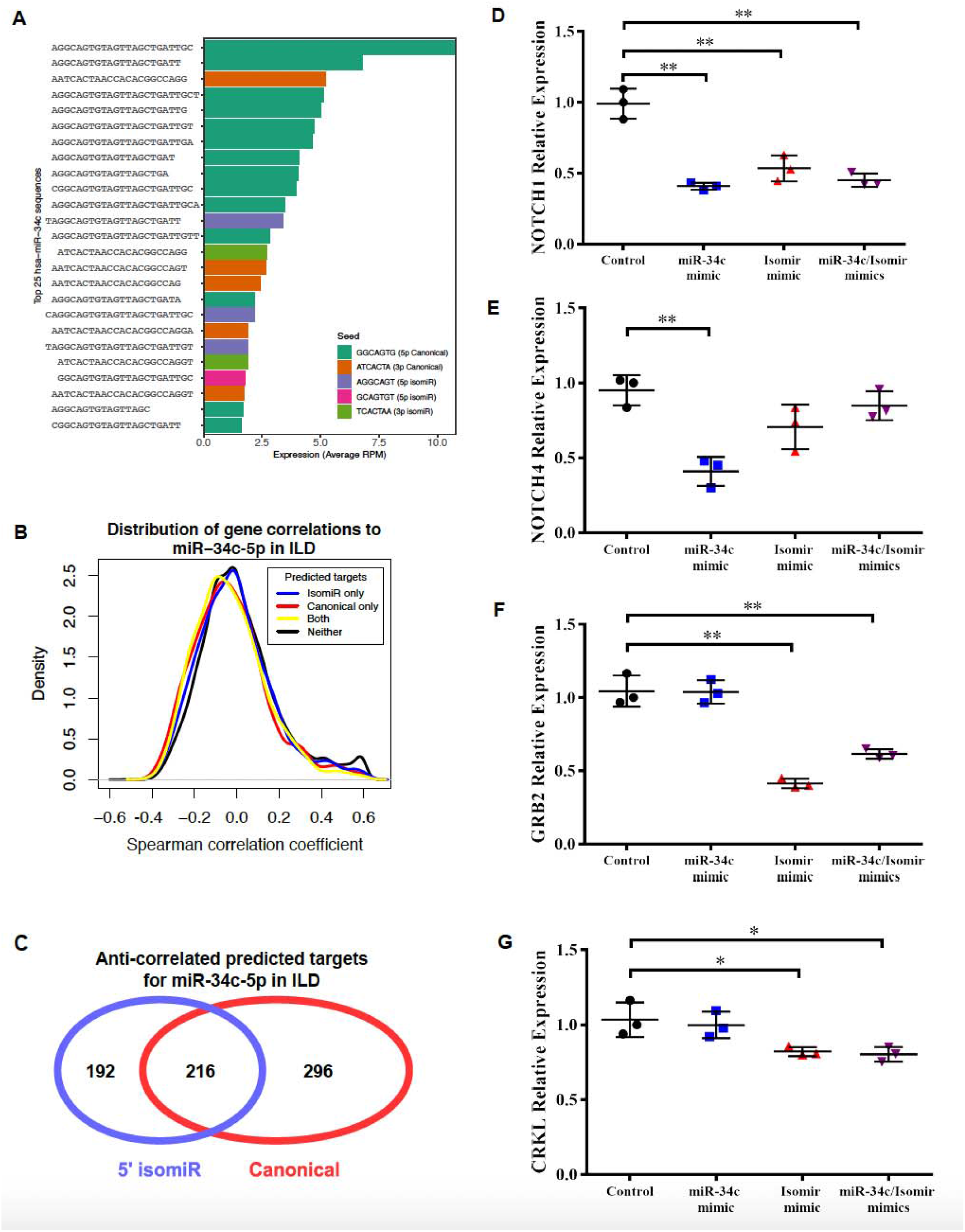
Functional roles of an isomiR for hsa-miR-34c. **A**. The top 25 highest expressed sequences are shown for the hsa-miR-34c locus. Three of these sequences represented an isomiR on the 5’ end which contained a non-canonical miRNA seed (purple). **B**. 5’ UTR-site predicted mRNA targets by Targetscan are more negatively correlated with the miR-34c isomiR seed than non-predicted targets (Kolmogorov-Smirnov test, p<1e-7). **C**. The overlap of negatively correlated (FDR<0.25) and 5’ UTR-site predicted targets of miR-34c and 5’ isomiR. miR-34c targets are significantly enriched for Notch signaling pathway by Enrichr (p<0.02); 5’ isomiR targets are significantly enriched for Ras signaling pathway by Enrichr (p<0.002). **D**. IMR90 fibroblast cells show significant repression of NOTCH1 with all experimental transfections (p<0.005, p<0.005, p<0.005). **E**. NOTCH4 expression is only downregulated by miR-34c mimic transfection (p<0.005) and not by transfection of the isomiR. **F**. GRB2 is significantly downregulated with the miR-34c 5’ isomiR mimic transfection (p<0.005, p<0.005) but not the miR-34c canonical transfection. **G**. CRKL is also significantly downregulated with the miR-34c 5’ isomiR mimic transfection (p<0.05, p<0.05) but not the miR-34c canonical transfection.

To explore similarities and differences in putative mRNA targets between canonical and isomiR seeds of miR-34c-5p, Targetscan v6.0^41^ was used to predict mRNA targets for each seed. mRNAs were grouped into four categories: predicted targets of the canonical and 5’ isomiR seeds, predicted targets of the canonical seed only, predicted targets of the the 5’ isomiR seed only, or those not predicted to be a target of either seed. Additionally, the expression of each gene was correlated to the overall expression levels of miR-34c-5p using Spearman correlation within the ILD samples. The distribution of correlation coefficients for groups of genes that were predicted targets of the canonical and/or 5’ isomiR seeds were more negative compared to non-predicted targets (Kolmogorov-Smirnov test; p < 1e-7; **Figure 4B**), suggesting that both the canonical seed and the 5’ isomiR seed may be negatively regulating target gene expression. We also explored the degree of overlap between genes that were significantly negatively correlated to miR-34c-5p expression (Spearman correlation; FDR q-value < 0.25) and were also a predicted target of either the canonical or 5’ isomiR (**Figure 4C**). Interestingly, 47% of the miR-34c-5p 5’ isomiR targets were distinct from the miR-34c-5p canonical targets. Using Enrichr,^53^ we found that the anti-correlated predicted targets specific to the 5’ isomiR were enriched for genes in the “Ras protein signal transduction” pathway including GRAP, GRB2, YWHAB, RHOA, RAPGEF6, MAPKAPK3, RALA, and SHC3 (p = 0.0001; **Supplementary Table 9**). Anti-correlated predicted targets specific to the miR-34c-5p canonical seed were enriched for the “notch signaling pathway” which contained other Notch-related genes beyond NOTCH1 including ADAM10, PSEN1, HEY1, DLL4, and NOTCH4 (p = 0.003 **Supplementary Table 10**). The predicted targets specific to the canonical seed were also enriched in other Ras-related pathways such as “Ras GTPase binding” and “small GTPase binding” suggesting that the canonical seed and the isomiR seed may be regulating different members of the same signaling pathway.

### Validation of expression and miR-34c-5p isomiR activity

The expression of miR-34c-5p was measured in a subset of ILD and Control samples (n = 10 per group) via qRT-PCR and was significantly upregulated with disease (p<0.05, **Supplementary Table 11, Supplementary Figure 6**). Similarly, NOTCH1 which is a known target of the canonical miR-34c-5p^54,55^ and a predicted target of the 5’ isomiR, was validated to be down-regulated in ILD samples compared to Controls when measured by qRT-PCR (p < 0.05; **Supplementary Figure 7**). Finally, qRT-PCR was used to measure the expression of genes in Ras signaling pathway that were anti-correlated predicted targets of the canonical seed (CRK) or the 5’ isomiR seed (RALA, GRB2, GRAP, RHOA, ARAP2, CRKL). Five of the seven predicted targets were significantly down-regulated in the subset of ILD samples compared to Controls (p < 0.05; **Supplementary Figure 7**). Additionally, two genes not predicted to be targets of miR-34c-5p seed were examined. An association between RHOC expression and ILD was observed with the mRNA microarrays (p = 0.002) and confirmed with qRT-PCR (p < 0.01) while a lack of association between EGF expression and ILD was observed with the mRNA microarrays (p = 0.230) and confirmed with qRT-PCR (p > 0.05; **Supplementary Figure 7**). These results confirm that associations with ILD determined by mRNA microarrays are largely recapitulated by qRT-PCR. Given the differences in the sets of predicted target genes for each miR-34c-5p seed sequence, we next sought to validate activity of the miR-34c 5’ isomiR. Human lung fibroblasts (IMR90) were transfected with mimics of miR-34c-5p, the miR-34c-5p 5’ isomiR, or both sequences. In the ILD cohort, NOTCH1, a known target of miR-34c-5p, was significantly anti-correlated with miR-34c-5p expression (FDR q-value = 0.016) and was a predicted target of the 5’ isomiR seed as well. Fibroblasts transfected with any mimics had significant down-regulation of NOTCH1 expression (p<0.005, **Figure 4D**). NOTCH4 is a validated target of the canonical form of miR-34c-5p,^55^ but was not a predicted targeted by the miR-34c-5p 5’ isomiR. Expression of NOTCH4 was only significantly decreased with canonical miR-34c-5p overexpression (p<0.005, **Figure 4E**). GRB2 and CRKL, genes involved in the Ras pathway and predicted targets of only the isomiR, were significantly downregulated only with the miR-34c-5p 5’ isomiR mimic transfections and not the canonical miR-34c-5p transfection (p<0.05, **Figure 4F**,**G**). Overall, these results demonstrate the ability of the miR-34c-5p isomiR to modulate the expression of predicted targets distinct from the canonical seed sequence.

## Discussion

We applied unsupervised and integrative analyses to multi-omic data to characterize the role of miRNAs in the setting of COPD and ILD. Novel subgroups of patients were identified with miRNAs that were differentially expressed in either disease. Several sample subgroups were enriched for disease patients and/or had significantly worse lung function phenotypes compared the subgroup with the most Control subjects (sample cluster S1). Interestingly, several disease patients were also in cluster S1. We have previously shown that both gene and miRNA expression can vary with regional emphysema severity across different sections within the lungs of patients with COPD^15^. Therefore, COPD or ILD patients clustering with Control patients could be due to variable sampling of diseased regions within the lung. Conversely, a smaller number of control patients clustered in one of the disease-associated subgroups. This could be due to the fact that in diseases such as COPD, aberrant processes like emphysema can begin to occur before the onset of overall lung function decline^56^. Overall, these molecular subgroups defined by miRNA expression represent previously unappreciated patient subclasses that may require distinct therapeutic modalities.

In order to identify potential miRNA regulators of gene expression, we leveraged genetic and gene expression data available on a subset of samples. We first identified eQTLs for both miRNAs and genes and found that, in contrast to gene expression, relatively fewer miRNAs were associated a *cis* SNP compared to protein-coding genes, suggesting that their regulation is more dependent on other factors within this disease setting. However, we were able to detect a large number of *trans* interactions, suggesting the lack of *cis* interactions is not simply due to lack of power. To identify potential miRNA-gene interactions within each disease, we applied the CIT and observed a significantly higher proportion of interactions for specific miRNA between disease and normal networks, including interactions for the miR-34 and miR-449 families. These miRNA families regulates mucociliary differentiation by directly targeting the NOTCH pathway^50,52,54,55,57^. Gene modules for these miRNAs in the COPD and ILD integrative networks were associated with airway epithelial cell differentiation in an independent dataset. Interestingly, miR-34b and miR-34c have been associated with emphysema severity^58^. We also found that the primate-specific miR-4423 was differentially connected in the COPD network. Expression of this miRNA is highly connected with the miR-449/34 family and has been previously associated with airway differentiation in smokers with lung cancer^45^. As the SNPs associated with the miR-449/34 family were on different chromosomes than the miRNAs, future studies will be needed to elucidate the mechanisms by which the trans-genetic variants can modulate the expression of these miRNAs.

Finally, we leveraged the ability of small-RNA sequencing to characterize sequence variation beyond expression levels and identified an isomiR with a novel seed sequence at the miR-34c-5p locus. This seed sequence was predicted to target a distinct set of genes from the canonical seed sequence and was enriched for genes involved in Ras signaling. This pathway has been previously implicated in tight junction formation in normal airway epithelial barrier formation.^59^ Down-regulation of this pathway may be necessary for normal differentiation of ciliary cells in the airway as well as the aberrant differentiation observed in a subset of ILD patients. Additional experiments in animal models will be necessary to determine if inhibition of these miRNAs can ameliorate disease phenotypes. The aggregation of these findings suggests a role for aberrant miRNA and isomiR regulation of airway differentiation in a subset of COPD or ILD patients and the inhibition of this process may represent a novel therapeutic approach for disease treatment.

## Data Availability

The microRNA expression datasets generated and analyzed during the current study are available in the Raw and normalized data is available at the Gene Expression Omnibus (GEO) under the accession number GSE201121.

## Acknowledgements

This work was supported by the National Institutes of Health/National Heart, Lung, and Blood Institute with funding from the Lung Genomics Research Consortium (RC2-HL101715) and R01HL118542 (M.E.L., A.S.).

## Author contributions

A.B.P., J.D.C. and J.B. contributed to data analysis. L.L., G.L., J.X., and Y.O.A. contributed to data generation. C.G. and D.T. performed experiments. B.J.G., J.T., I.V.Y., and F.S. contributed to sample collection and processing. A.B.P. and J.D.C. wrote the manuscript. M.W.G., D.A.S., N.K., A.S., and M.E.L. contributed to the overall study design. All authors reviewed the manuscript.

## Notes

### Competing Interest Statement

The authors have declared no competing interest.

