## Supplementary Figures for "Integrative genetic and genomic networks identify microRNA associated with COPD and ILD"

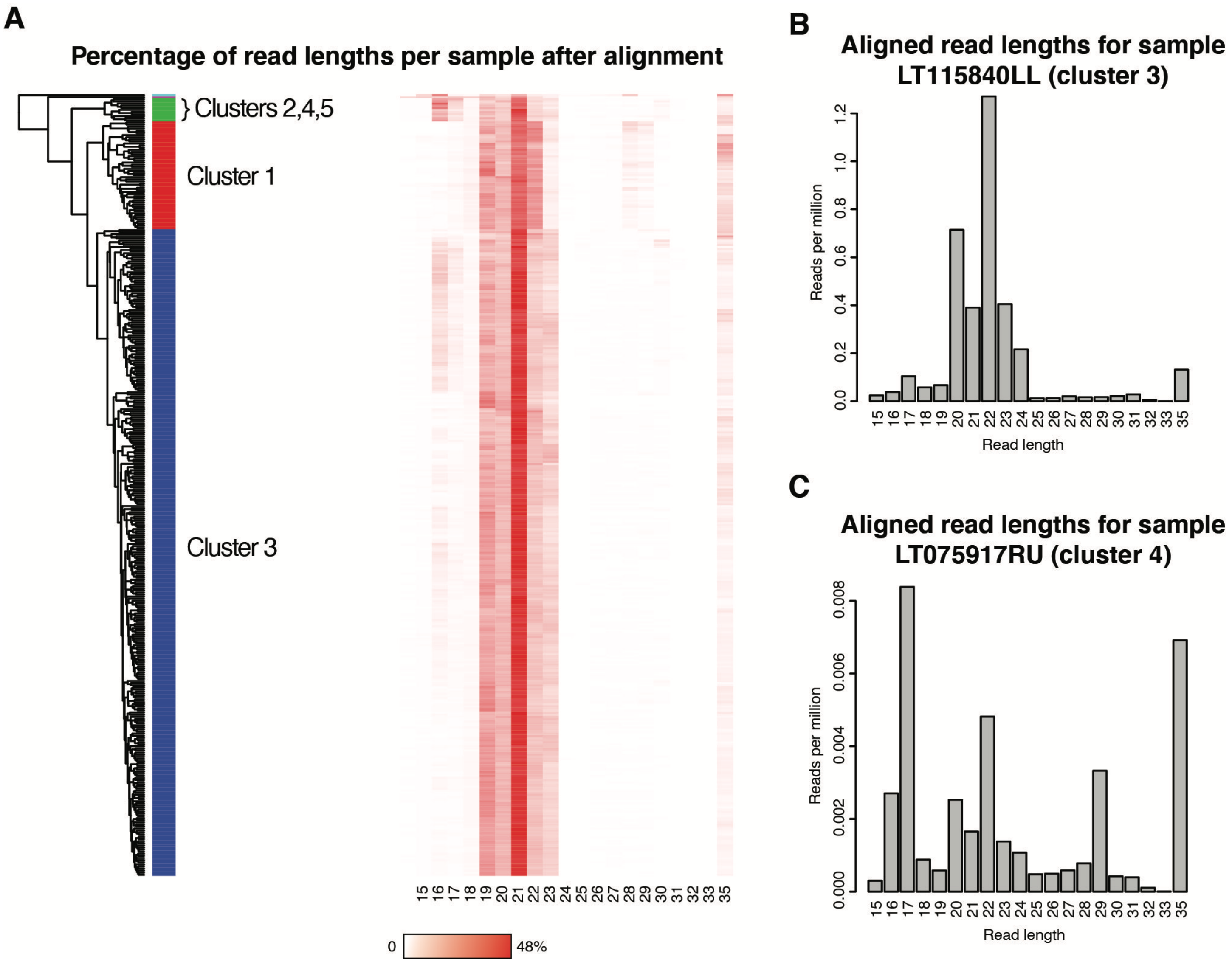


**Supplementary Figure 1. Sample filtering using the distribution of aligned read lengths.** The small RNA of 371 samples was sequenced and reads with a length less than 15 bases were excluded after trimming. **(A)** After alignment with Bowtie, samples were clustered on the normalized distribution of aligned read lengths using the Jensen-Shannon divergence as the distance metric and hierarchical clustering with average linkage. The 5 most distinct clusters were identified with the *cutree* function. The majority of samples fell into clusters 1 and 3. The distribution of aligned read lengths for samples in clusters 1 and 3 had a peak at 21 bases, which is expected as the small RNA fraction should largely contain miRNAs. Clusters 2, 4, and 5 had aligned read length distributions with peaks at other bases and were excluded from the down-stream analysis. **(B)** LT115840LL is a representative sample of cluster 3 which had expected read length distributions. **(C)** LT075917RU was in cluster 4 and had unexpected peaks at 17 and 29 bases and thus was excluded from the down-stream analyses.

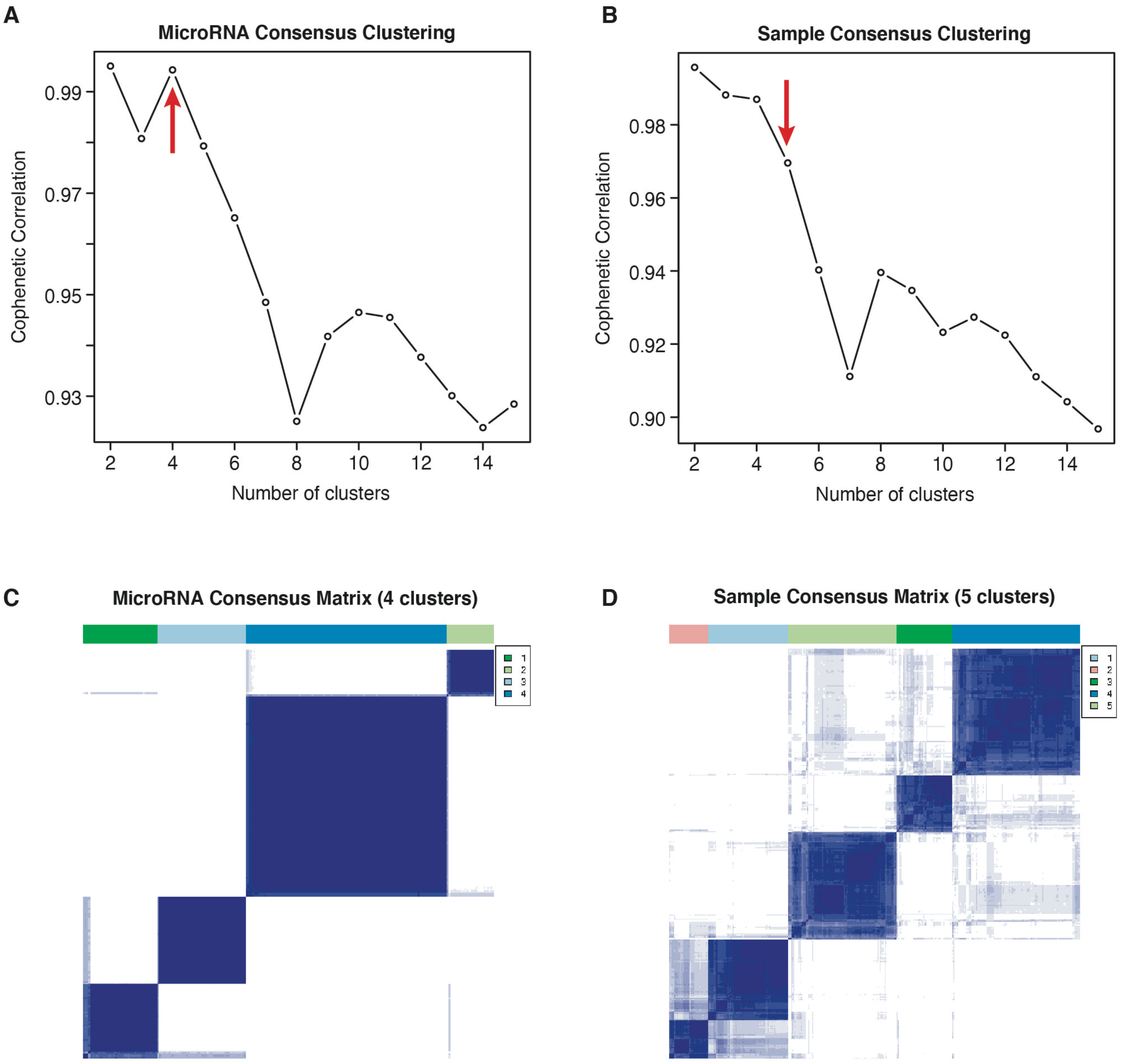


**Supplementary Figure 2. Consensus clustering of miRNAs and samples.** The *ConsensusClusterPlus* R package was used to separately cluster miRNAs and samples into group. The Cophenetic coefficient was used to help determine the choice of numbers of cluster. **(A)** Four was the most optimal number of clusters for the miRNA. **(B)** While the most optimal number of clusters was 2 for the samples, a large drop in the Cophenetic coefficient could be observed after the 5-cluster solution. The consensus heatmaps for the **(C)** miRNA and **(D)** samples show that the items largely clustered within their respective groups. The only exception was the sample clusters 1 and 2 which has some cross-clustering of samples. MicroRNA clusters 1, 2, 3, and 4 were renamed to m4, m2, m3, and m1, respectively, when displayed in the heatmap in **Figure 1**. Similarly, Samples clusters 1, 2, 3, 4, and 5 were renamed to s5, s4, s3, s1, and s2, respectively, when displayed in the heatmap in **Figure 1**.

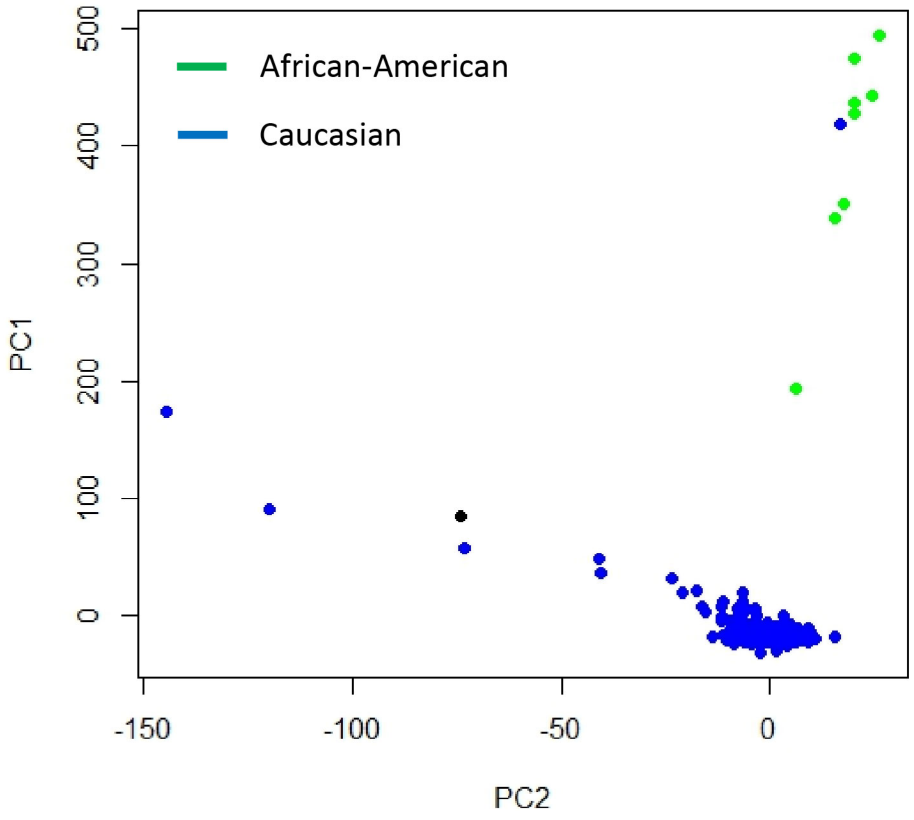


**Supplementary Figure 3. PCA plot showing the genetic population structure measured with the SNP chips.** Points are labeled by self-reported ethnicity.

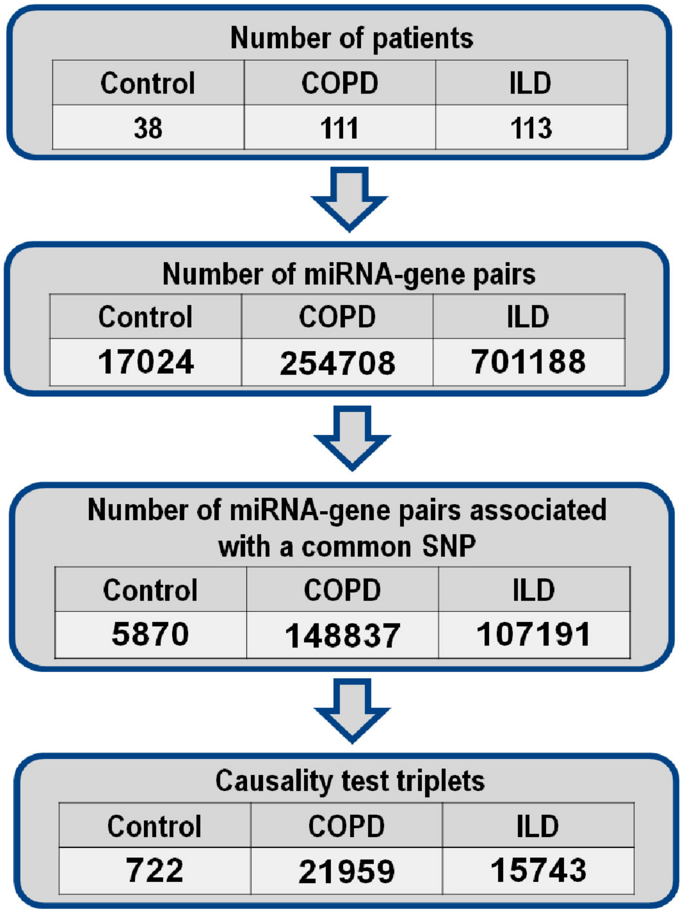


**Supplementary Figure 4. CIT network construction.** Number of significant interactions at each step of network construction in COPD, ILD and control groups.

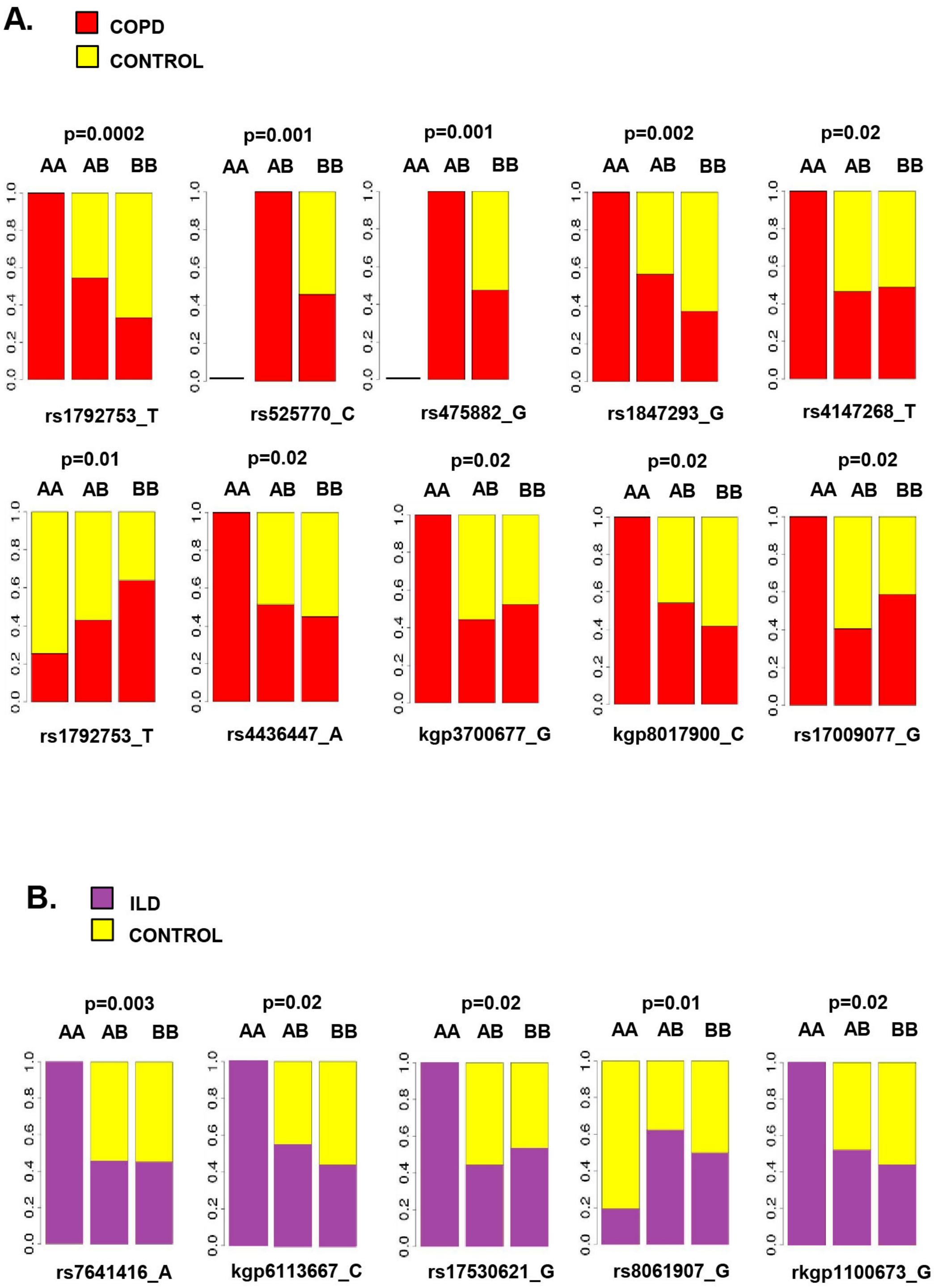


**Supplementary Figure 5. A.** SNPs that are significantly associated with COPD by a Fisher's exact test (q<0.25). **B.** SNPs that are significantly associated with ILD by a Fisher's exact test (q<0.25).

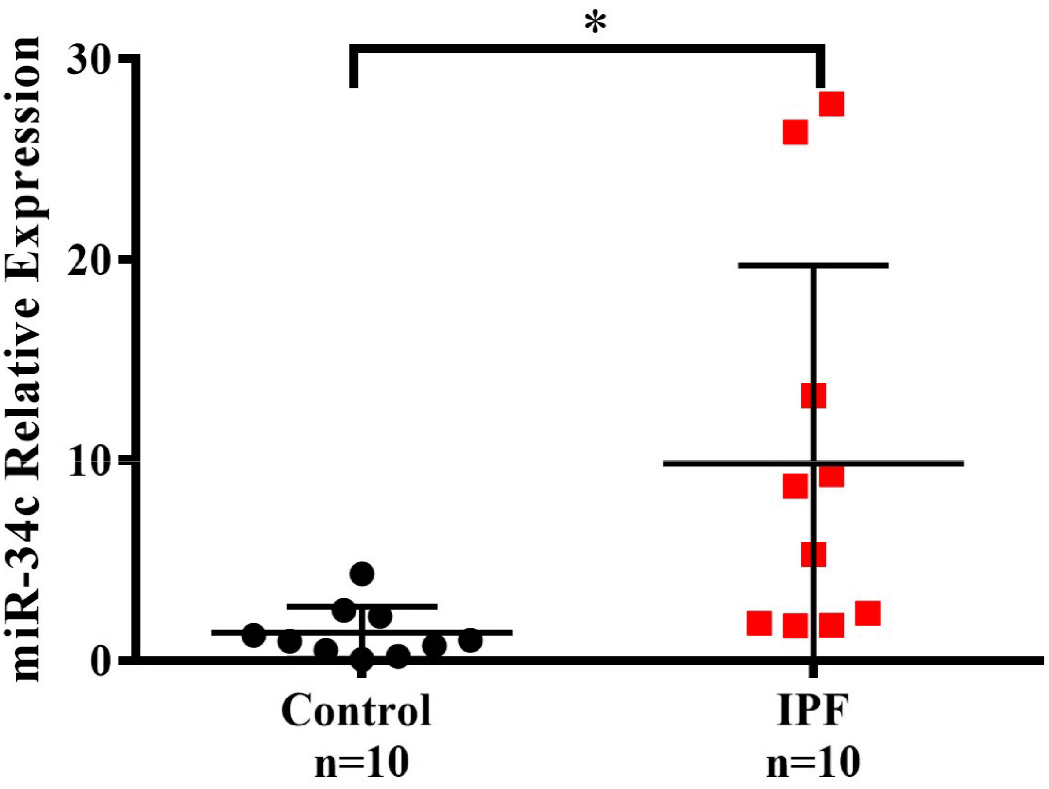


**Supplementary Figure 6.** Upregulation of miR-34c in ILD tissue validates by qRT-PCR. IPF (n=10) tissues have a significantly higher expression of miR-34c then control (n=10) tissues ($p<0.05$).

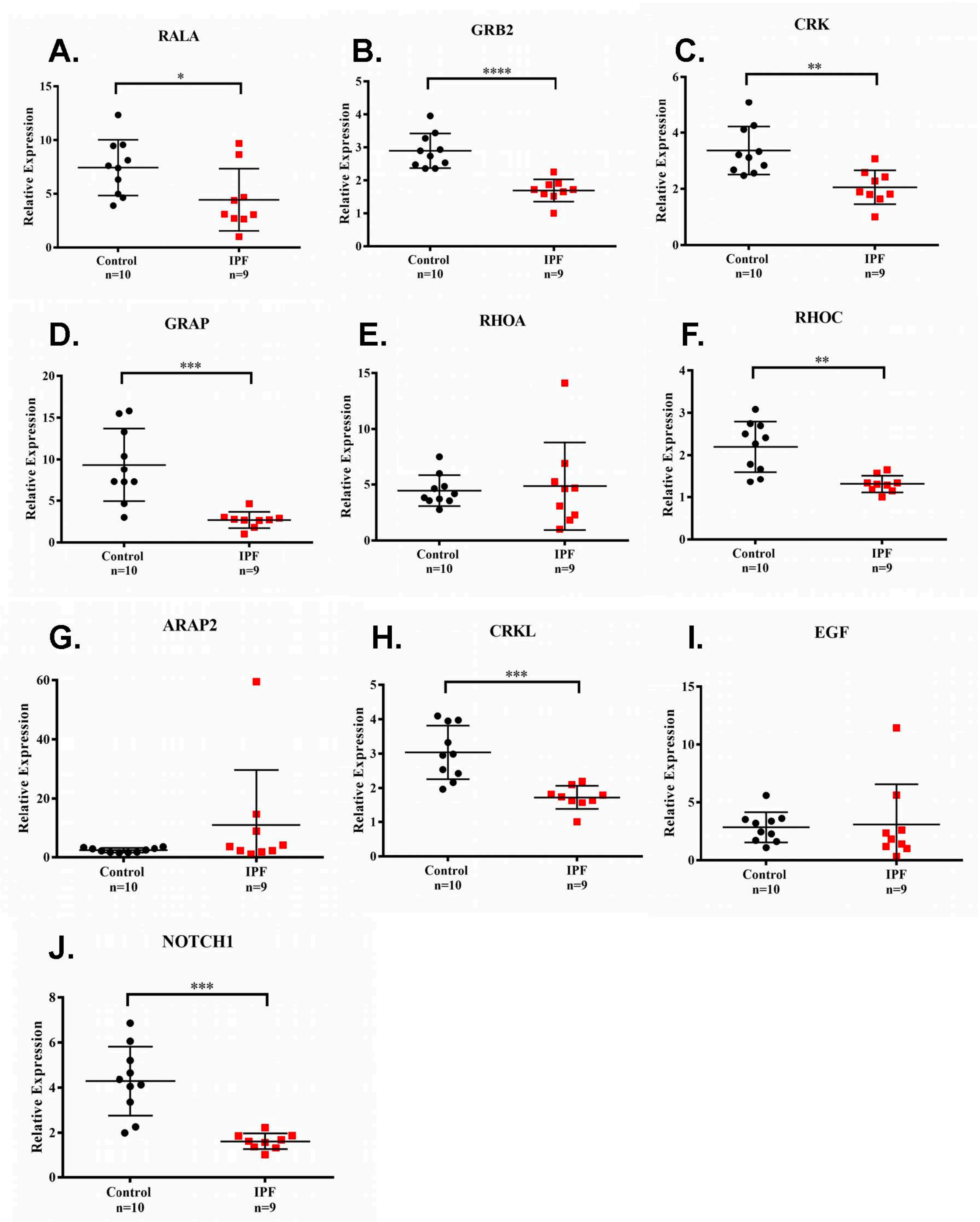


**Supplementary Figure 7.** Of the ten genes measured in clinical samples, nine of which are members of the Ras protein signaling pathway, seven validated as being downregulated in ILD compared to control tissue. These seven genes are: **A**. RALA ($p<0.05$), **B**. GRB2 ($p<0.00005$), **C**. CRK ($p<0.005$), **D**. GRAP ($p<0.0005$), **F**. RHOC ($p<0.005$), **H**. CRKL ($p<0.0005$), and **J**. NOTCH1 ($p<0.0005$).
